# “Knowledge and practices towards COVID-19 during its outbreak: a multinational cross-sectional study”

**DOI:** 10.1101/2020.04.13.20063560

**Authors:** Abdallah Y Naser, Eman Zmaily Dahmash, Hassan Alwafi, Zahra Khalil Alsairafi, Ahmed M. Al Rajeh, Yosra J Alhartani, Fawaz Mohammad Turkistani, Hamad S. Alyami

## Abstract

**Background:** The emergence of COVID-19 globally coupled with its unknown aetiology and its high transmission rate has created an unprecedented state of emergency worldwide. Public knowledge and awareness about COVID-19 are essential in suppressing its pandemic status.

**Method:** A cross-sectional study using an online survey was conducted between 19th of March and 6th of April 2020 in three Middle Eastern countries (Jordan, Saudi Arabia and Kuwait) to explore the knowledge and practices of Middle Eastern population towards COVID-19. A previously developed questionnaire was used. Multiple linear regression analysis was used to identify predictors of COVID-19 knowledge.

**Results:** A total of 1,208 participants were involved in this study from the three countries (Jordan = 389, Saudi Arabia = 433, and Kuwait = 386). The majority of participants (n = 810, 67.2%) were females and aged 30 to 49 years (n = 501, 41.5%). Participants had moderate overall COVID-19 knowledge with a mean score of 7.93 (±1.72) out of 12, 66.1%. Participants had better knowledge about disease prevention and control with 83.0%, whereas the lowest sub-scale scores were for questions about disease transmission routes (43.3%). High education level was an important predictor of greater COVID-19 knowledge scores (p<0.01).

**Conclusion:** Middle Eastern participants are of a relatively low level of knowledge about COVID-19, particularly regarding its transmission routes. Policymakers are recommended to develop informative COVID-19 related campaigns targeted specifically towards university students, unemployed individuals and those with lower levels of education.

## Introduction

Coronavirus disease 2019 (abbreviated “COVID-19”) is an infectious disease with unknown aetiology characterised with acute pneumonia that has been recognized in Wuhan, China in December 2019. Predominantly, it is characterised by fever, fatigue, and dry cough. Approximately, 20.0% of COVID-19 patients developed to the severe stage, which is characterised by acute respiratory distress syndrome, bleeding, and coagulation dysfunction (1). In the Middle East region, this virus has appeared in some countries before others. For example, it has been firstly identified in Iran then spread in Kuwait, Saudi Arabia, and Jordan. The spread of the virus in Kuwait earlier than others could be attributed to the fact that there was a national holiday in Kuwait in the period between the 23^rd^ to the 29^th^ of February, where most people used to spend it abroad. As people returned from vacations, confirmed cases with COVID-19 started to raise. In March 12, 2020, the World Health Organization (WHO) declared that the COVID-19 outbreak is a pandemic (2). The pandemic lingers to take a hefty toll on healthcare professionals, families, communities and the whole world. In response to that, Jordan, Saudi Arabia, and neighbouring Kuwait took drastic measures early on in a bid to contain the COVID-19 pandemic, which includes halting air travels, imposing curfews, and quarantining and testing thousands of individuals (3, 4).

Once a new pandemic starts, the focus of research and action within the medical and public health communities is chiefly and rightly directed towards the identification of the cause, clinical presentation, diagnosis as well as treatment (5). Few studies will address the epidemiology of the disease, the effectiveness of preventive measures, and the population psycho-behavioural directions. Nevertheless, addressing public health preventive measures deserve equal attention. Although there are several ongoing clinical trials assessing potential vaccines and treatments for COVID-19, currently there is no specific treatment that can combat the virus within medical practice. Therefore, public health measures are of crucial value. Such measures mandate the public to be aware of transmission means and have safe preventive practices (6). In this regard, people had concerns about the safety measures to be taken in order to protect themselves and their families from being infected. Generally, the spread of any infectious disease is associated with a high level of fear among the population (7). A particular concern in this regard is the spread of misinformation about COVID-19 on social media sites (8). Universal data collection and analysis of pandemic control hardly contains information about the knowledge and practices of the population regarding the disease and their significance (9, 10). To guarantee the best control over the transmission of the disease, community’s adherence to important control measures are vital, which is affected to large extent by their knowledge and practices towards COVID-19 (9, 10).

Public practices that are built on knowledge ought to be evaluated during pandemics as it will enable policymakers to unravel generic issues from culture specific concerns and then communicate the best practices that appear to have an impact in successfully controlling the outbreak across different communities (11, 12). Thus there is an urgent need to understand what people knows about COVID-19, and which misperceptions they hold about this condition, in order to help healthcare authorities and the media to design effective information campaigns, to know which group of people should be targeted, and to help controlling the spread of the virus among population. Therefore, this study aims to assess the knowledge and practices of Middle Eastern population towards COVID-19 during its rapid rise period.

## Methods

### Study design and study population

A cross-sectional study by means of online survey was conducted between 19^th^ of March and 06^th^ of April 2020 in three Arab countries (Jordan, Saudi Arabia and Kuwait) to explore the knowledge and practices of Middle Eastern population towards COVID-19.

### Sampling strategy

A convenience sample of eligible participants was invited to participate in the study from the three countries through social media (Facebook and WhatsApp). All participants voluntarily participated in the study and were thus considered exempt from written informed consent. Study aim and objectives were clearly explained at the beginning of the survey. The inclusion criterion was participants aged 18 years who was living either in Jordan, Saudi Arabia or Kuwait. Participants were excluded if they were: a) below 18 years of age; or b) unable to understand Arabic language.

### Questionnaire tool

A previously developed questionnaire by Zhong et al. was adapted and used in this study (1). The original questionnaire was developed based on the National Health Commission of the People’s republic of China’s guidelines for clinical and community management of COVID-19 (13, 14). The original questionnaire had 12-questions (true/false questions): four regarding clinical presentations (K1 - K4), three regarding transmission routes (K5 - K7), and five regarding prevention and control (K8 - K12) of COVID-19. These questions were answered on a true/false basis with an additional “I don’t know” option. A correct answer was given a score of one point, and any false or unknown answer was given a score of zero point. The total knowledge score could range from 0 to 12, with a higher score indicating a better knowledge about COVID-19. Cronbach’s alpha coefficient of the knowledge questionnaire was 0.7 in our sample, showing acceptable internal consistency. Respondents’ practices were assessed using two behaviour questions, which asked the participants about whether they have gone to any crowded place, and wearing a mask when going out in recent days. In addition, the following information was collected about participants’ demographics: age, gender, marital status, education level, income, and employment status.

### Sample size

Using a confidence interval of 95%, a standard deviation of 0.5, a margin of error of 5%, the required sample size was 385 participants from each study population.

### Ethical approval

This study was approved by the Research Ethics Committee at Faculty of Pharmacy in Isra University, Amman, Jordan. As participation in the study was voluntary, the Research Ethics Committee approved consent waiver.

### Statistical analysis

Descriptive statistics were used to describe participants’ demographic characteristics. Continuous data were reported as mean ± SD. Categorical data were reported as percentages (frequencies). Independent samples t-test/one-way analysis of variance (ANOVA) was used to compare the mean knowledge scores between different demographic groups. Participants’ scores were interpreted as a continuous scale based on the scale midpoint, where scores above the midpoint identified stronger COVID-19 knowledge. Mean score was expressed in percentage out of 100% to facilitate the comparison between different sub-scales. Multivariable linear regression analysis was used to identify predictors of knowledge score. A two-sided p<0.05 was considered as statistically significant. The statistical analyses were carried out using SPSS (version 25).

## Results

### Participants’ characteristics

A total of 1,208 participants took part in this study (Jordan = 389, Saudi Arabia = 433, and Kuwait = 386). **Table 1** details the baseline characteristics of the participants in the three countries.

**Table 1:** Participants characteristics from each country.

| **Demographics** | **Overall**<br>**(n = 1,208)** | **Jordan**<br>**(n = 389)** | **Saudi Arabia**<br>**(n = 433)** | **Kuwait**<br>**(n = 386)** |
| --- | --- | --- | --- | --- |
| **Gender** (n= 1,205) No. (%) |  |  |  |  |
| Female | 810 (67.2) | 280 (72.0) | 324 (75.0) | 206 (53.6) |
| **Age** (n= 1,206) No. (%) |  |  |  |  |
| 18 – 29 years | 424 (35.2) | 237 (61.1) | 131 (30.3) | 56 (14.5) |
| 30 – 49 years | 501 (41.5) | 112 (28.9) | 189 (43.6) | 200 (51.9) |
| 50 years and above | 281 (23.3) | 39 (10.1) | 113 (26.1) | 129 (33.5) |
| **Marital status** (n= 1,207) No. (%) |  |  |  |  |
| Single | 429 (35.5) | 213 (54.8) | 148 (34.2) | 68 (17.7) |
| Married | 685 (56.8) | 161 (41.4) | 241 (55.7) | 283 (73.5) |
| Divorced | 57 (4.7) | 5 (1.3) | 31 (7.2) | 21 (5.5) |
| Widowed | 36 (3.0) | 10 (2.6) | 13 (3.0) | 13 (3.4) |
| **Education level** (n= 1,198) No. (%) |  |  |  |  |
| Completed secondary grade or lower | 274 (22.9) | 48 (12.5) | 105 (24.3) | 121 (31.7) |
| Completed bachelor degree | 723 (60.4) | 283 (73.7) | 244 (56.5) | 196 (51.3) |
| Completed higher education | 201 (16.7) | 53 (13.8) | 83 (19.2) | 65 (17.0) |
| **Employment status** (n= 1,206) No. (%) |  |  |  |  |
| Retired | 158 (13.1) | 13 (3.3) | 63 (14.5) | 82 (21.4) |
| Unemployed | 278 (23.1) | 113 (29.0) | 116 (26.8) | 49 (12.8) |
| Employed | 568 (47.1) | 147 (37.8) | 185 (42.7) | 236 (61.5) |
| University students | 202 (16.7) | 116 (29.8) | 69 (15.9) | 17 (4.4) |
| **Income** (n= 1,115) No. (%) |  |  |  |  |
| 705 \$ or lower | 368 (33.0) | 200 (58.3) | 130 (32.5) | 38 (10.2) |
| 705 \$ – 1,408 \$ | 175 (15.7) | 96 (28.0) | 62 (15.5) | 17 (4.6) |
| 1,408 \$ – 2,113 \$ | 75 (6.7) | 15 (4.4) | 26 (6.5) | 34 (9.1) |
| 2,113\$ and above | 497 (44.6) | 32 (9.3) | 182 (45.5) | 283 (76.1) |
| **In recent days, have you gone to any crowded place?** (n= 1,194) No. (%) |  |  |  |  |
| Yes | 183 (15.3) | 85 (21.9) | 38 (8.9) | 60 (16.0) |
| **In recent days, have you worn a mask when leaving home?** (n= 1,195) No. (%) |  |  |  |  |
| Yes | 605 (50.1) | 204 (52.6) | 191 (44.3) | 210 (55.9) |

The majority of participants (n = 810, 67.2%) were females, aged 30 to 49 years (n = 501, 41.5%), married (n = 685, 56.8%), has bachelor degree (n = 723, 60.4%), employed (n = 568, 47.1%), and with an income of 2,113$ and above (n = 497, 44.6%). Around 15.3% (n = 183) of the participants reported that they have gone to crowded places recently. Besides, around 50.1% (n = 605) of the participants reported that they wore mask when they leave their home.

**Table 2** below shows the mean scores (and score out of 100%) for COVID-19 knowledge questionnaire per subscale. The overall COVID-19 knowledge score was 66.1%. Participants had better knowledge about disease prevention and control with 83.0%, whereas the lowest sub-scale scores were for questions about disease transmission routes (43.3%).

**Table 2:**
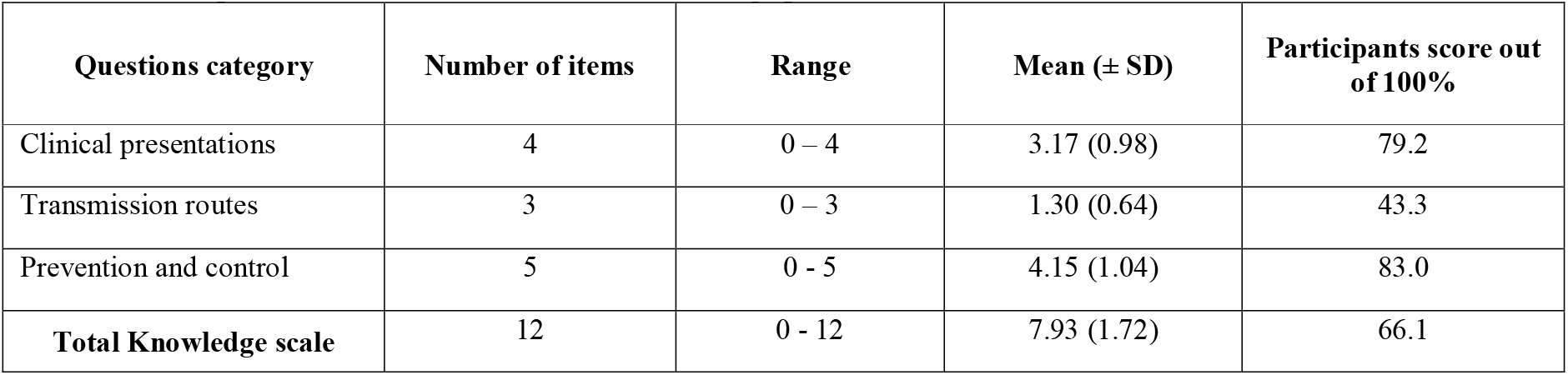
Participants mean scores for COVID-19 knowledge per subscale.

**Table 3** below details the correct answer rates of the 12 questions on the COVID-19 knowledge questionnaire.

**Table 3:** The correct answer rates of the COVID-19 knowledge questionnaire.

| Questions | % |
| --- | --- |
| K11. Isolation and treatment of people who are infected with the COVID-19 virus are effective ways to reduce the spread of the virus. | 98.4 |
| K10. To prevent the infection by COVID-19, individuals should avoid going to crowded places such as train stations and avoid taking public transportations. | 97.9 |
| K12. People who have contact with someone infected with the COVID-19 virus should be immediately isolated in a proper place. In general, the observation period is 14 days. | 95.1 |
| K7. The COVID-19 virus spreads via respiratory droplets of infected individuals. | 92.1 |
| K1. The main clinical symptoms of COVID-19 are fever, fatigue, dry cough, and myalgia. | 91.5 |
| K3. There currently is no effective cure for COVID-2019, but early symptomatic and supportive treatment can help most patients recover from the infection. | 89.3 |
| K4. Not all persons with COVID-2019 will develop to severe cases. Only those who are elderly, have chronic illnesses, and are obese are more likely to be severe cases. | 69.5 |
| K2. Unlike the common cold, stuffy nose, runny nose, and sneezing are less common in persons infected with the COVID-19 virus. | 68.7 |
| K8. Ordinary participants can wear general medical masks to prevent the infection by the COVID-19 virus. | 57.0 |
| K5. Eating or contacting wild animals would result in the infection by the COVID-19 virus. | 30.3 |
| K6. Persons with COVID-2019 cannot infect the virus to others when a fever is not present. | 9.2 |
| K9. It is not necessary for children and young adults to take measures to prevent the infection by the COVID-19 virus. | 4.1 |

Participants had the highest rates of correct answers for questions related to the necessity of isolation and some procedures related to prevention and control of the infection (K10 – K12). On the other hand, the lowest rates of correct answers were for questions related to susceptibility of children and young adults to COVID-19, transferability of the infection from asymptomatic patients, and eating habits related to COVID-19 transmission (K5, K6, and K9).

### Participants’ demographics and COVID-19 knowledge

**Table 4** below presents participants’ demographics data and their COVID-19 knowledge scores. Participants’ knowledge scores significantly differed by country, age, marital status, education level, and whether they wear mask upon leaving home or not (p*<*0.05).

**Table 4:** COVID-19 knowledge score by participants characteristics (n = 1,208).

| COVID-19 knowledge score |  |  |  |
| --- | --- | --- | --- |
| Variable | Mean | SD | P-value |
| **Country** |  |  |  |
| Jordan | 8.44 | 1.46 | 0.000*** |
| Saudi Arabia | 7.79 | 1.57 |  |
| Kuwait | 7.59 | 1.99 |  |
| **Gender** |  |  |  |
| Males | 7.86 | 1.94 | 0.263 |
| Females | 7.97 | 1.61 |  |
| **Age** |  |  |  |
| 18 – 29 years | 7.72 | 2.04 | 0.006** |
| 30 – 49 years | 8.01 | 1.49 |  |
| 50 years and above | 8.11 | 1.55 |  |
| **Marital status** |  |  |  |
| Single | 7.75 | 2.07 | 0.002** |
| Married | 8.07 | 1.46 |  |
| Divorced | 7.47 | 1.72 |  |
| Widowed | 8.19 | 1.56 |  |
| **Education level** |  |  |  |
| Completed secondary grade or lower | 7.72 | 2.01 | 0.015* |
| Completed bachelor degree | 7.94 | 1.70 |  |
| Completed higher education | 8.18 | 1.31 |  |
| **Employment status** |  |  |  |
| Retired | 7.82 | 2.02 | 0.092 |
| Unemployed | 7.97 | 1.78 |  |
| Employed | 8.04 | 1.51 |  |
| University students | 7.70 | 1.91 |  |
| **Income** |  |  |  |
| 705 \$ or lower | 8.01 | 1.70 | 0.094 |
| 705 \$ – 1,408 \$ | 8.24 | 1.69 | |
| 1,408 \$ – 2,113 \$ | 7.81 | 1.65 | |
| 2,113\$ and above | 7.92 | 1.38 | |
| **In recent days, have you gone to any crowded place?** |  |  |  |
| Yes | 8.11 | 1.89 | 0.325 |
| No | 7.99 | 1.49 |  |
| **In recent days, have you worn a mask when leaving home?** |  |  |  |
| Yes | 8.16 | 1.51 | 0.001** |
| No | 7.86 | 1.59 |  |

\* $p < 0.05$ , \*\* $p < 0.01$ , \*\*\* $p < 0.001$

Multiple linear regression analysis showed that individuals who had a bachelor degree or higher education level had a greater knowledge scores (p*<*0.01). On the other hand, divorced individuals, university students, individuals with an income 1,408 $ and above had a lower knowledge score (p*<*0.05) **Table 5**.

**Table 1:** Multiple regression analysis predicting participants’ COVID-19 knowledge.

|  | Model 1<sup>a</sup> |  |  |
| --- | --- | --- | --- |
| **Variable** | **B** | **SE** | **β** |
| **Demographic data** |  |  |  |
| **Age** |  |  |  |
| 30 – 49 years | 0.162 | 0.154 | 0.051 |
| 50 years and above | 0.343 | 0.197 | 0.093 |
| **Gender** |  |  |  |
| Females | 0.047 | 0.102 | 0.014 |
| **Marital status** |  |  |  |
| Married | 0.059 | 0.138 | 0.019 |
| Divorced | -0.644 | 0.246 | -0.089** |
| Widowed | 0.050 | 0.324 | 0.005 |
| **Educational level** |  |  |  |
| Completed bachelor degree | 0.329 | 0.123 | 0.103** |
| Completed higher education | 0.519 | 0.159 | 0.127** |
| **Employment status** |  |  |  |
| Unemployed | -0.263 | 0.207 | -0.067 |
| Employed | -0.138 | 0.180 | -0.044 |
| University students | -0.582 | 0.241 | -0.137* |
| **Income** |  |  |  |
| 705 \$ – 1,408 \$ | -0.014 | 0.152 | -0.003 |
| 1,408 \$ – 2,113 \$ | -0.456 | 0.213 | -0.073* |
| 2,113\$ and above | -0.547 | 0.152 | -0.174*** |
| Constant |  |  | **7.967** |
| Adjusted R<sup>2</sup> |  |  | **0.029** |
| P-value |  |  | **P<0.001** |

\*p < 0.05, \*\*p < 0.01, \*\*\*p < 0.001
a: includes age, gender, marital status, educational level, employment status, and income level
B: the average change in the dependent variable associated with a 1 unit change in the independent variable, statistically controlling for the other independent variables; SE: it is the standard deviation of its sampling distribution or an estimate of that standard deviation; β: a statistical measure that compares the strength of the effect of each individual independent variable to the dependent variable

## Discussion

To the best of our knowledge, this is the first study that assesses the knowledge and practices towards COVID-19 among the Arabic speaking Middle Eastern countries. The results revealed that participants are still embracing misconceptions about COVID-19, resulting in an insufficient practice of protective measures against COVID-19 infection. However, some of those individuals who did not adhere to protective measures could be the one who were in duties that necessitate being in a crowded places such as port employees and working in the healthcare sector. The findings of this study revealed that the overall knowledge score among the three countries was around 66.1% (score 7.93 out of 12) with the highest was among Jordanians with a mean score of (8.44 (±1.46); 70.3%) indicate an average level of knowledge about the pandemic. Among the three countries, the participants achieved the lowest knowledge score in the “transmission routes” sub-scale with a score of 43.3%. Whereas the highest knowledge score was recorded on the “preventive and control measures” sub-scale with a score of 83.0%. Overall, the total knowledge score (66.1%) was moderate, which was unexpected as this epidemiological survey was conducted during the advanced stage of the pandemic.

Although the low percentage 15.3% (183/1,194) with regard to the reported practices of attending crowded places were encouraging, only 50.1% (605/1,195) of participants declared that they wore masks when leaving home. The World Health Organization (WHO) during Feb 2020 indicated that there is a priority for research to focus on actions that can save lives now which included *“Optimise use of protective equipment and other infection prevention and control measures in health care and community settings – It is critical to protect health care workers and the community from the transmission and create a safe working environment”* (10). The needs to avoid the crowd and wear masks upon leaving the house are key practices and top priority of the WHO. A similar study conducted by Zhong et.al. (2020) in China (1), showed that knowledge score was 90.0% and citizens were compliant with protective measures, where 96.4% avoided crowded places and 98.0% wore masks upon departing their homes during COVID-19 outbreak (1). These results undoubtedly revealed that there were significant gaps in the public knowledge about COVID-19 within the Arabic speaking Middle Eastern countries and hence suggest the need for improving individuals’ COVID-19 knowledge via health education, which may cause improvements in their practices towards COVID-19 (1).

Although the situation of the pandemic is severe and the news reports on this public health emergency through various channels of information and via hotlines of the Ministries of Health, a lot of people learn about this infectious disease from invalid social media resources such as WhatsApp, Instagram, Twitter, etc. This has also been mentioned by Gelsetzer (2020) (15), where it has been noticed that a substantial proportion of people in the United States (US) and the United Kingdom (UK) believe in the “myth busters” who continue to circulate inaccurate information about COVID-19. Such information included that rinsing nose with saline, using a hand dryer, taking antibiotics, and gargling mouthwash could be effective preventive measures against COVID-19, and that receiving a letter or package from China may pose a risk of Covid-19 infection. Other unreliable beliefs of the general public in both the US and the UK about COVID-19 were that it is a fatal disease and that children are at high risk of death from it. Another disbelief was that medical masks are highly effective in protecting from catching a COVID-19 infection (15). However, people in the Middle East region have reported some similar beliefs and other different. For example, in the current study, most participant (66.9%) reported that it is unnecessary for children and younger adults to take preventive measures against COVID-19. Thus, in contrast to people in the US and UK, people in the Middle East believe that children and younger adults are at lower risk of COVID-19 infection. On the other hand, most people (57.0%) in the Middle East shared similar beliefs regarding wearing medical masks to prevent them from COVID-19 infection. Regarding the fatality feature of COVID-19, Middle East participants reported conflicting beliefs to people in the US and UK, as the majority (89.0%) believe that although there is no effective treatment, supportive measurements and symptoms alleviation can cure the infection.

Unfortunately, the present study showed that 15.3% of participants still go to crowded places and around 50.0% do not wear masks when leaving homes recently. However, not wearing a mask when leaving home could be related to that most of the participants (84.7%) were very cautious and avoided crowded places. This strict preventive practice from the participants could be primarily attributed to the very strict prevention and control measures implemented by local governments such as traffic limits all throughout the countries and the shutdown of shopping malls and work in all sectors except the health sector. Second, this could be due to the population fear of the virus and its high infectivity. In China, although a study by Zhong et al. (2020) was conducted during the very early stage of the pandemic, people achieved a correct rate of 90.0% on COVID-19 knowledge questionnaire (1). In addition, the population’s adherence to the preventive and control measures was higher than in the Middle East, where almost all people stayed home during the virus outbreak period, and wear masks when leaving homes. This could be attributed to the more serious situation of the COVID-19 pandemic in China, where higher rates of death were recorded, resulting in higher beliefs and adherence to preventive measures.

Further analysis of the findings was directed towards determining demographic factors associated with knowledge gaps among participants. Such findings will be valuable for public-health policymakers and healthcare professionals to recognize target populations for health education activities on the COVID-19 outbreak. This is critical as the countries involved in the study are still at the beginning of the outbreak and governments took extreme measures to contain the pandemic. But such measures, which include imposing a curfew, are virtually useless unless were accompanied by responsible individuals’ preventative practices (3). In this study as well as in previous studies in China, US, and UK, an association has been noticed between particular demographic characteristics and knowledge, beliefs, and practices towards COVID-19 (1, 15). For example, in China, an association between non-adherence to preventive and control measures and male gender, occupation of “students”, marital status of “others”, residing in other parts of China, and poor COVID-19 knowledge has been reported (1). Those findings were supported by previous studies regarding age and gender patterns of risk-taking behaviours (16, 17). In the US and UK, Geldsetzer (2020) has reported lower fatality rates due to COVID-19 infection among individuals of East-Asian ethnicity and children (15). In the current study, there was a significant difference between knowledge scores in the three countries included in the analyses, and the highest knowledge score among the participants has been found in Jordan. Both males and females showed similar levels of knowledge about COVID-19.

Higher COVID-19 knowledge scores were found to be significantly associated with age and educational attainment, which is in line with the study conducted in China during the COVID-19 pandemic (1). Older individuals above the age of 29 years showed a significant increase in knowledge scores than younger ones. Similarly, an appreciable increase in knowledge score with the increase in education level was noted. This could suggest that the contents and forms of health-related information about the pandemic could not be understandable and acceptable to young adults and the less educated individuals (12). Our findings are consistent with several previously published studies that related age and the level of education with knowledge and awareness about outbreaks (18, 19). Furthermore, several studies reported that the lack of knowledge contributed to the emergence and spread of the outbreak. Therefore, providing targeted health education programs to raise awareness is vital. Such programs need to be tailored to young individuals and particularly those with lower education levels (19-21).

Furthermore, a significant negative effect on the knowledge score was the employment status, specifically among students (p<0.05). The results are aligned to the finding that lack of knowledge is related to young people (18 – 29 years). Such findings are alarming, as the lack of knowledge about COVID-19 is correlated with a higher prevalence rate of the infection (1, 22). Therefore, there is a need for improving students’ knowledge about COVID-19 by means of health education, which may also result in improvements in their practices towards COVID-19. Such results could support the decision made by many countries including Jordan, Kuwait, and Saudi Arabia in taking a political commitment to temporally close all educational institutions to contain the spread of the COVID-19 pandemic. Globally, the action involved 188 countries, affecting around 1.5 billion learners. The mode of teaching moved into emergency remote learning (22, 23). Yet, although the closure of schools and educational facilities is a good mean to prevent young adults from mixing with others, it will not prevent exposure even with curfew laws. Young adults will always go out and get exposed to the risk of getting infected as the level of knowledge is low. A recent study on COVID-19 concluded that educational institutions closures alone would preclude only 2–4% of deaths, which is a lot less than other social distancing interventions. The integration of additional social distancing strategies along with educational institutions closures needs to be considered (24). Furthermore, the results of our study pertinent to knowledge scores stratification according to demographics are in line with studies on COVID-19 in China and severe acute respiratory syndrome (SARS) outbreaks among others (1, 25). These findings also proposed that health education strategies would be more effective if it was designed to targets certain demographic groups, such as university students.

Good knowledge is vital to enable individuals to have better practices in pandemics and outbreaks. A study on SARS demonstrated that a higher knowledge score was found to be connected with improved adherence to precautionary practices (11, 26). Of interest, an income of ≥1,409 $ (p<0.05) had a significant negative association with the knowledge score in our cohort, showing that high income does not improve knowledge and practices. However, this is contrary to a previous study on the influenza pandemic, where there was no association between knowledge and income (26). A possible reason for such result is that the higher income of individuals is accompanied with low education level. The current study showed that knowledge scores significantly differed across marital status categories (P<0.002), divorced participants had a significant negative association with COVID-19 knowledge score (p<0.01). Our findings were in line with the results obtained from the study on COVID-19 in China, which revealed a statistically significant difference in knowledge score according to marital status, however, they found that the category denotes as others (included divorced and widowed, separated, remarried) showing the highest score in knowledge (11.0 out of 12.0) (91.7%), P <0.001) (1). This difference could be justified by two elements. Firstly, different countries might have different cultural dimension and hence results could not be compared. Secondly, the group used in the Chinese study included all divorced, separated, widowed and remarried individuals and hence we could not build a good correlation (1).

Overall, knowledge scores within our cohort were significantly associated with a higher likelihood of dangerous practices towards COVID-19 pandemic. Such findings indicate the presence of knowledge gap and the need to improve individuals’ COVID-19 knowledge via extensive health education strategies. Such strategies are to be tailored and targeted towards specific demographic groups particularly, university students, individuals with low education, higher income and divorced.

### Strengths and limitations

To the best of our knowledge this is the first study in the Arabic Speaking Middle Eastern countries that investigated the public knowledge and practices during COVID-19 pandemic. A strength of the study is that a large sample of participants were recruited during this critical period - the actual COVID-19 outbreak - and hence replies reflect the actual status. The participants were from three countries namely, Jordan, Saudi Arabia and Kuwait, which increases the generalisability of these findings. Additionally, the use of previously used assessment tool that allowed comparison with other population was another strength of the study. However, there are some limitations. The study design itself, a cross-sectional survey design, limited our ability to identify causality between study variables. There are limited studies that assessed knowledge and practices of individuals during COVID-19 pandemic worldwide and in the Middle East specifically, which limited our ability to compare our findings with Arabic-speaking countries of a similar socioeconomic level and culture. In this study we employed a quantitative methodology with pre-set responses, which might not have allowed participants’ views to provide varied but useful qualitative information. In addition, we used an online survey for data collection and therefore, some vulnerable populations within the three countries under the COVID-19 pandemic could not be reached and we may have missed some of the targeted population. However, we tackled this by distributing the survey among three different countries and widely used the social media. Finally, we were not able to estimate the response rate for our questionnaire study.

## Conclusion

Findings of this study suggest that Middle Eastern participants are of a relatively low level of knowledge about COVID-19, particularly regarding its transmission routes. The majority of the population showed appropriate practice by avoiding crowded places and staying home during the rapid rise period of the COVID-19 outbreak. However, not wearing a mask when leaving home was predominant. As good COVID-19 knowledge is associated with optimistic attitudes and appropriate practices towards COVID-19, policymakers are in a position to develop targeted in information campaigns provided to targeted population specifically university students, unemployed individuals and those with lower levels of education. This information could be conveyed to people by clinicians to their patients, and news coverage supplied by the media and social media platforms.

## Data Availability

The data that support the findings of this study are available from the corresponding author upon reasonable request.

## Acknowledgments

This study was supported by Isra University (Amman, Jordan) and Najran University and Cancer Society in Najran, Saudi Arabia.

## Author contributions

AN conceived the study, wrote the methods, conducted the formal analysis, and coordinated the study. AN, EZD, ZA, and AA drafted the manuscript with input from all authors. All authors have been involved in drafting the manuscript or revising it critically for important intellectual content. All authors read and approved the final manuscript.

